# The mutational spectrum of Jervell and Lange-Nielsen syndrome: insights from highly consanguineous families

**DOI:** 10.1101/2025.10.22.25336867

**Authors:** Aya Hataba, Mona Allouba, Mariam Fathy, Alaa Afify, Eslam Ahmed, Mohamed Riad, Mohamed Elmaghawry, Youssef El Bayoumy, Amany Elleithy, Aliaa Mahfouz, Jodie Ingles, Aya Galal, Sohila Rabie, Mohamed Allam, Sarah Halawa, Nour Elsadek, Besra Samy, Heba Kassem, Omnia Kamel, Yasmine Aguib, Magdi H. Yacoub

## Abstract

**Background/Objectives:** Jervell and Lange-Nielsen syndrome (JLNS) is an autosomal recessive disease caused by mutations in *KCNQ1* or *KCNE1*. It is characterized by prolonged QT interval on electrocardiogram, deafness and an increased risk of sudden cardiac death (SCD) (25%). Given the high consanguinity rate (35%) in the Middle East and North Africa region (MENA), an enrichment of JLNS is expected, providing a powerful opportunity to identify ancestry-specific genetic determinants. We report genetic data from the largest MENA JLNS cohort analyzed to date.

**Methods:** Prospective Egyptian JLNS families and ancestry-matched controls were recruited to Aswan Heart Centre for clinical phenotyping and genetic testing for genes with reported roles in inherited cardiac conditions (Illumina). Variants were classified according to the ACMG/AMP guidelines.

**Results:** We followed up 57 patients (25 males and 32 females; mean age at enrollment: 5.8 years) from 47 families for four years. Consanguinity was reported in 76.2% of families with complete family data. All of the sequenced patients (n=52) harbored homozygous variants in *KCNQ1* (n=51) or *KCNE1* (n=1). Among these variants, 90.4% were truncating. The majority of KCNQ1 variants were located in the c-terminal domain. No significant differences in the clinical parameters were observed between carriers of different domains of the KCNQ1 protein, as well as between carriers of truncating variants versus nontruncating variants. A novel, recurring *KCNQ1* variant (c.912G>A) was identified in 11 Egyptian patients from 10 families (21.2%), suggesting that it could be an Egyptian-specific founder mutation.

**Conclusion:** Variants discovered in this underrepresented population add to the mutational spectrum of JLNS and uncover ancestry-specific genetic determinants of disease. Expanding the cohort will provide important genetic and mechanistic insights into the global understanding of inherited arrhythmia syndromes.

## Introduction

Ion channelopathies is a group of hereditary electrical disorders of the heart that manifest despite the existence of a normal cardiovascular anatomy^1^. They are characterized by being lethal, rare, and are difficult to diagnose^1^. One example of an ion channelopathy is Long QT Syndrome (LQTS). It is characterized by prolongation of the QT interval on electrocardiogram (ECG) along with ventricular arrhythmias (VAs)^1,2^. According to the 2022 ESC guidelines for the management of patients with ventricular arrhythmias and the prevention of sudden cardiac death (SCD), the diagnostic criteria for LQTS include obtaining a score of greater than 3 on the ‘Schwartz’ scoring system^3,4^ a QTc that is greater than or equal to 480 ms^2^.LQTS may be inherited in an autosomal dominant or an autosomal recessive manner with or without other non-cardiovascular manifestations^2,5,6^. This paper will focus on the autosomal recessive form of LQTS with additional non-cardiac manifestation entitled Jervell and Lange-Nielsen Syndrome (JLNS).

JLNS is a cardio-auditory syndrome, or Surdo-Cardiac Syndrome, that is characterized by prolongation of the QT interval on ECG and bilateral sensorineural deafness that manifests at birth^7–10^. As a recessive disease, it has a globally variable prevalence that is estimated around 3-5 patients per 1,000,000^11–13^. An additional manifestation includes syncope that is triggered by emotion, physical effort, or physical activity^9,11^. Cardiac manifestations in patients diagnosed with JLNS typically appear in half of the patients by approximately 3 years old, and in the majority by 18 years old^9,14^. Thus, this renders the patients with an increased risk of SCD^7^.

The genes implicated in causing JLNS are *KCNQ1* and *KCNE1*, in which homozygous or compound heterozygous mutations may lead to the disease^9,15–21^. In symptomatic patients, the majority of JLNS-causal mutations occur in the *KCNQ1* gene, and those with KCNQ1 mutations are at a higher risk of experiencing arrhythmic events^9^. On the other hand, in asymptomatic patients, the majority of JLNS-causal mutations are harbored on the *KCNE1* gene^9^.

To date, no studies have investigated the genetic aetiology of JLNS in patients of Egyptian ancestry. Nonetheless, the genetic etiology of JLNS has been investigated through small-scale studies, including case-studies, in several countries of the Middle East and North Africa (MENA) region, including Morocco^15,22,23^, Turkey^15,21,24,25^, Algeria^15^, Saudi Arabia^26–28^, and Iran^29–31^. Given the high consanguinity rate (35%) in the Egyptian population and the associated increase in homozygosity, a higher prevalence of JLNS is expected compared to outbred populations of European ancestry. This study aims to define the genetic architecture of JLNS among patients of Egyptian descent, validate the findings published to date regarding the genetics of JLNS and expand the mutational spectrum of JLNS.

## Methods

### Study Cohorts

#### 1. Egyptian JLNS Cohort

A prospective cohort of JLNS patients (n=57), denoted hereafter as Egypt JLNS, was recruited to the specialized inherited arrhythmia clinic at Aswan Heart Center (AHC). As a tertiary centre, the patients were referred from other facilities or diagnosed during active screening of families of probands. Clinical diagnosis was confirmed through electrophysiological evaluation at AHC’s outpatient clinic through dedicated clinical form. Twelve lead ECG was obtained for the patients and their parents. QT interval was measured from start of QRS complex to the end of T wave and then corrected to the heart rate using Bazett formula. Diagnosis of JLNS was made in patients with prolonged QTc along with presence of sensorineural hearing loss. The Egypt JLNS cohort includes families with more than one JLNS patient.

#### 2. Egyptian Control Cohort

Egyptian healthy volunteers (n = 400), denoted hereafter as Egypt controls, were recruited from across the country as part of the ECCO-GEN. All volunteers underwent detailed phenotyping, including clinical examination, 12-lead electrocardiogram, and CMR to confirm the absence of cardiovascular disease. Principal component analysis (PCA) showed that both Egypt HCM patients and controls overlapped into one distinct cluster, confirming that they are ancestry-matched^32,33^.

Participants from both cohorts provided informed consent and were approved by their local research ethics committees.

### Gene Selection

DNA samples of both Egypt JLNS cohort and Egypt Controls were sequenced on Illumina MiSeq and NextSeq 550 platforms using the TruSight Cardio Sequencing Panel for inherited cardiac conditions (ICC)^34^. Analysis was focused on the two causative genes of JLNS, which are *KCNQ1* and *KCNE1*.

### Rare Variants Classification

Variants were annotated by CardioClassifier based on rules defined by the American College of Medical Genetics and Genomics/Association for Molecular Pathology (ACMG/AMP)^35,36^. Rare variants in *KCNQ1* or *KCNE1* were defined as having a filtering allele frequency (FAF) of (8.2 × 10^−6^) for LQTS^37^. However, it is important to note that these databases underrepresent individuals of Middle Eastern and North African (MENA) ancestry, which may lead to inaccurate classification of variant rarity—i.e., variants classified as ‘rare’ in global datasets could, in fact, be more common in under-sampled populations..though noting that Middle eastern and North African ancestries are not well represented in existing population reference databases.

### Statistical analysis and visualization

Statistical analysis was performed using the framework applied by Morgat and colleagues (2024) in their recent work on the characterization of *KCNQ1* variants^38^. Following their example, we used the Shapiro–Wilk test to assess the distribution of continuous variables. For normally distributed data, we utilized the Welch Two Sample t-test to compare the means between two groups, the one-way ANOVA (Analysis of Variance) Test to compare the means between more than two groups, and the Tukey HSD (Honestly Significant Difference) post-hoc test to identify the significantly different groups from each other. For non-normally distributed data, we utilized the Kruskal–Wallis test. For categorical dependent variables, we used the Fisher’s exact test. Means ± standard deviation were used to showcase continuous variables, whereas frequency and percentage were used to present categorical variables. Unless Bonferroni Correction is applied, statistical significance was determined for a p-value less than 0.05.

In order to define the genetic architecture of JLNS in patients of Egyptian descent, homozygous rare variants identified in *KCNQ1* in JLNS patients were selected for burden testing using Fisher’s exact test against the ancestry-matched Egyptian control cohort. Boneferroni correction was applied based on the number of tested variants to account for multiple testing. Further, the distribution of the variants by protein domain was compared between both study cohorts to evaluate if rare variants are particularly enriched in certain clusters across the *KCNQ1* protein and the effect of localization on clinical parameters. Statistical analysis and visualization was performed using R (4.4.0).

## Results

### Egypt JLNS Cohort characteristics

The Egyptian JLNS cohort comprises a total of 57 JLNS patients (43.9% males and 56.1% females) from 47 families who were followed up for a mean duration of four years (Table 1). The mean age at first event was approximately three years old in patients with complete phenotypic data (39/57). All patients displayed prolonged QT interval on ECG along with congenital bilateral sensorineural deafness. The majority of patients were symptomatic, experiencing syncope (74.5%) and sudden cardiac arrest (3.6%; n=2); whereas, 14 patients (25%) were totally asymptomatic (Table 1). Family history of SCD was reported in half of the cohort. Similarly, family history of other channelopathy was described in almost 50% of the cohort. Consanguinity (defined as first- or second-cousin marriage) was reported in 76.2% of families with complete family data (32/42). A total of five patients (8.8%) have experienced SCD (Table 1).

**Table 1:** Demographic and cardiac characteristics of Egypt JLNS cohort.

| General characteristics | Egypt JLNS Cohort |
| --- | --- |
| Demographics |  |
| Males n(%) | 25 (43.9%) |
| Females n(%) | 32 (56.1%) |
| Mean Age at Enrollment (sd) | 5.8 ($\pm 5.7$) |
| Range of Age at Enrollment | 0 – 28 |
| Mean Age at First Event (sd) | 3.102564 ($\pm 2.5$) |
| Range of Age at First Event | 0 – 15 years |
| Consanguinity no. of families (%) | 32/42 families (76.2%) |
| Family history of SCD n(%) | 29 (50.9%) |
| Family history of other inherited channelopathy n(%) | 27/56 (48.2%) |
| **Cardiac characteristics** |  |
| Average QTc Measurement (sd) | 544.386 ($\pm 55.37526$) |
| Range of QTc Measurement | 400 – 705 ms |
| Deafness n(%) | 57 (100%) |
| Syncope n(%) | 41/55 (74.5%) |
| Resuscitated Sudden Cardiac Arrest (SCA) n(%) | 2/55 (3.6%) |
| SCD n(%) | 5 (8.8%) |
| Asymptomatic n(%) | 14/56 (25%) |
| Number (%) of Families with History of SCD History | 26/47 (55.3%) |
| ICD Implantation | 7/56 (12.5%) |
| Cochlear Device Implantation | 25/55 (45.5%) |
| Mean Hemoglobin (sd) | 10.12 ($\pm 1.8$) (g/dl) |
| Range of Hemoglobin | 4.5 – 13.6 (g/dl) |

### Electrocardiographic phenotype of Egyptian JLNS patients and healthy parents

After confirming the normal distribution of QTc measurements, we assessed whether sex and disease groups (JLNS patient vs parent) were associated with statistically significant differences in QTc. Consequently, we obtained the following categories: female JLNS patient (n=32); male JLNS patient (n=25); female parent of JLNS Patients (n=30); and male parent of JLNS Patients (n=25). JLNS patients, regardless of their reported gender, had significantly longer QTc measurement than their parents (Figure 1, Supplementary Table 1). Specific details of the electrocardiographic phenotype of the four categories can be found in Supplementary Table 2. Further, 8.8% (5/57) of patients and 85.5% (47/55) of parents had a QTc measurement below 460 ms. In contrast, 47.4% (27/57) of patients had a very prolonged QTc measurement (>550 ms), while none of the parents had a QTc measurement exceeding this threshold.

**Figure 1:**
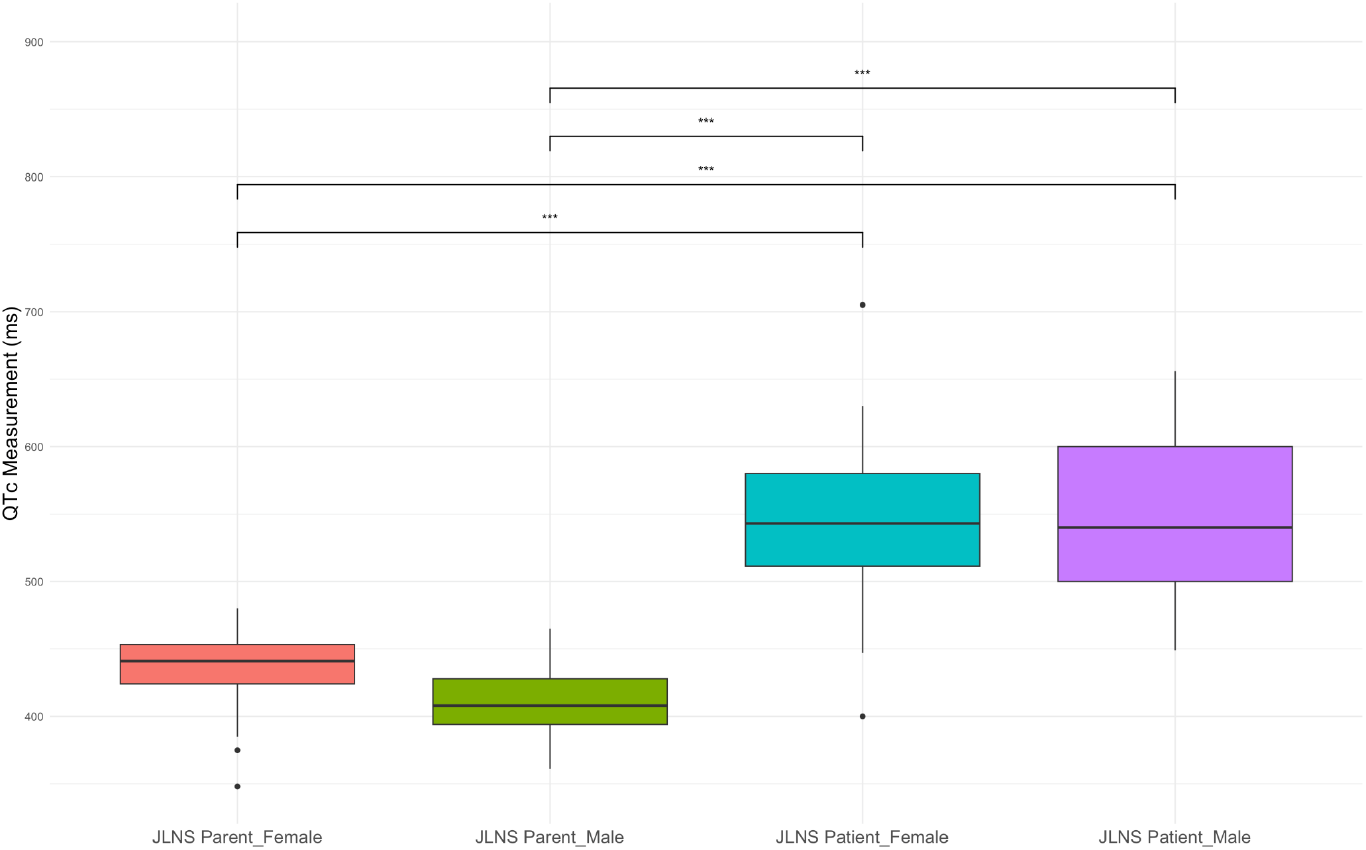
JLNS patients have significantly prolonged QTc measurements compared with their parents, irrespective of the reported sex. The box plots showcase the categorization of the electrocardiographic phenotype of JLNS patients versus that of their parents based on the reported sex. Comparisons were performed using the Tukey HSD Test, where stars of significance are shown above each significant comparison. *** represent p-values < 0.001.

Moreover, we evaluated the electrocardiographic phenotype of both symptomatic and asymptomatic Egypt JLNS cases. According to the Welch Two Sample t-test, there was no statistical significance in the difference between the QTc measurements of both groups (p-value = 0.33). Table 2 depicts the mean, standard deviation, range, and median of QTc measurements of symptomatic versus asymptomatic Egypt JLNS cases.

**Table 2:**
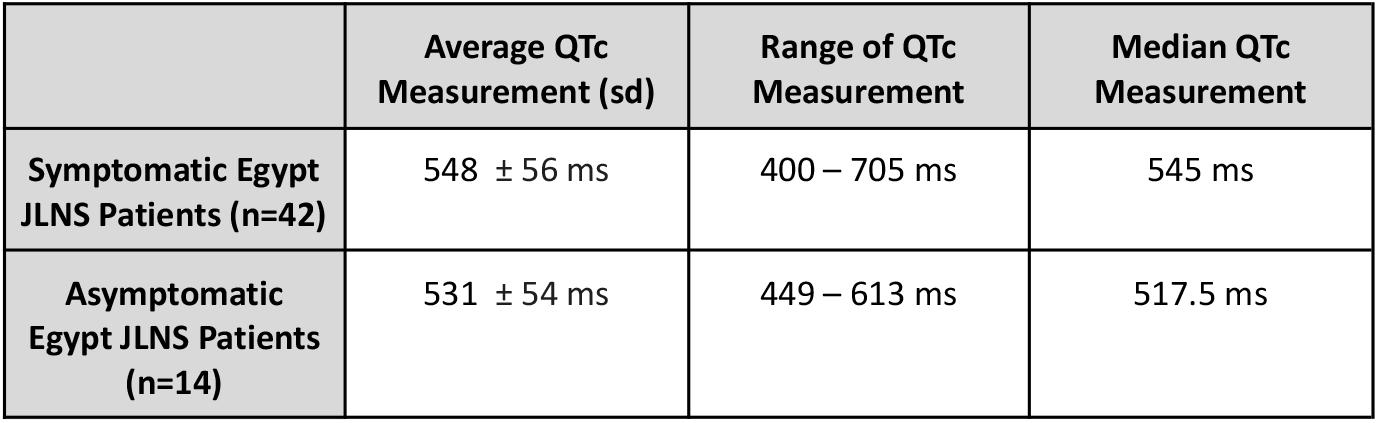
Electrocardiographic phenotype of symptomatic versus asymptomatic Egypt JLNS cases.

|  | Average QTc Measurement (sd) | Range of QTc Measurement | Median QTc Measurement |
| --- | --- | --- | --- |
| **Symptomatic Egypt JLNS Patients (n=42)** | 548 ± 56 ms | 400 – 705 ms | 545 ms |
| **Asymptomatic Egypt JLNS Patients (n=14)** | 531 ± 54 ms | 449 – 613 ms | 517.5 ms |

### Mutational spectrum of Egyptian JLNS patients

A total of 52 Egypt JLNS patients underwent targeted genetic testing for n=174 genes with reported roles in ICC (Illumina). Among those with sequencing data, 98% of patients harbored genetic variants in *KCNQ1* (51/52) (Figure 2). The remaining patient (n=1) harbored a homozygous, truncating variant of uncertain significance in *KCNE1*. No rare homozygous variants in *KCNQ1* or *KCNE1* were observed in the Egypt Control cohort.

**Figure 2:**
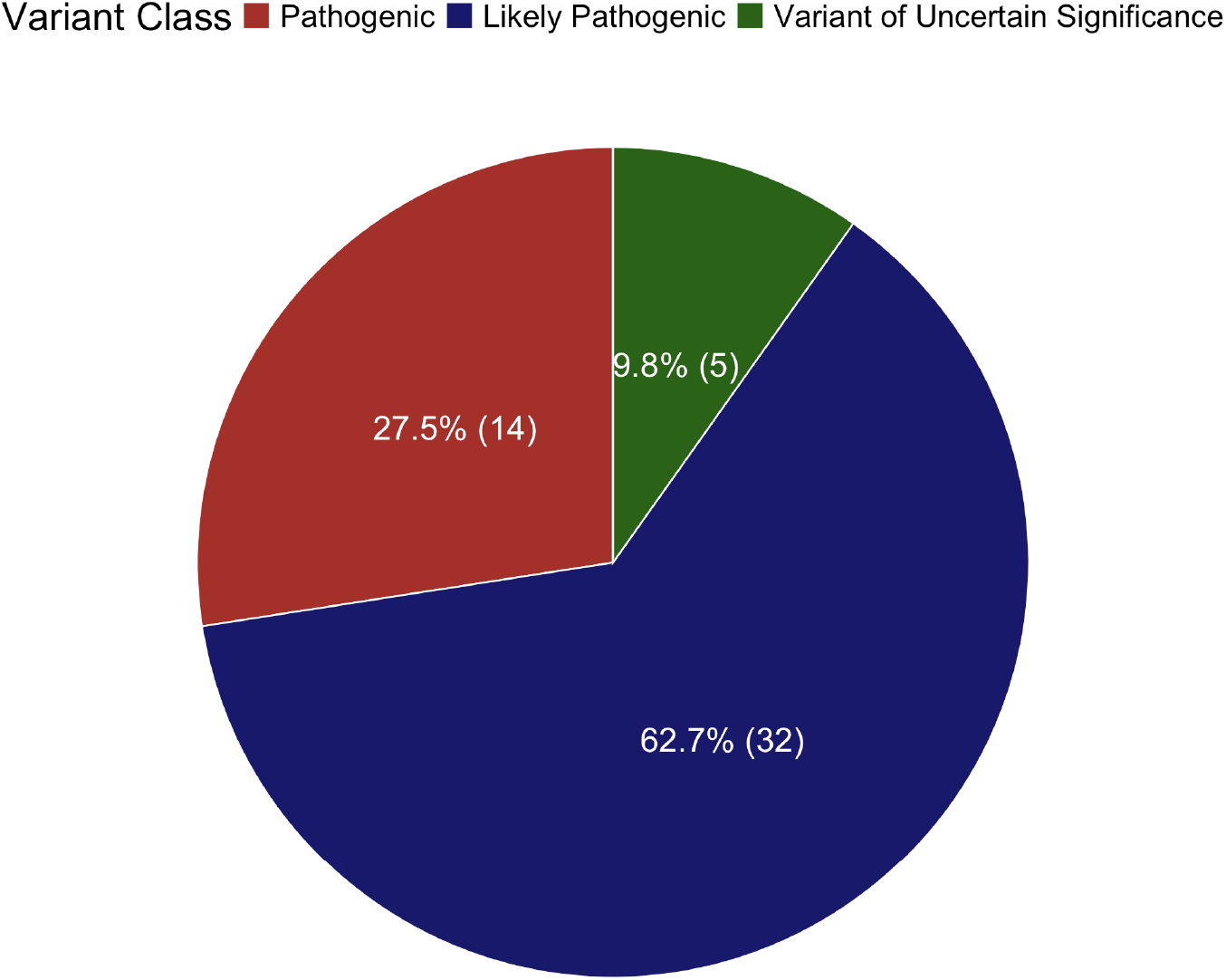
Distribution of Patients (n= 51) harboring genetic variants in *KCNQ1*, as per the ACMG/AMP variant classification.

The majority of homozygous variants identified in the Egypt JLNS cohort were truncating. A summary of the identified clinically actionable variants can be found in Table 6. Several recurrently observed homozygous variants, including p.Arg518Ter, p.Glu449ArgfsTer14, and p.Trp304Ter were significantly enriched in the Egypt JLNS populations compared to controls (26.92% vs 0, p-value=1.272e-14 for p.Arg518Ter and p.Glu449ArgfsTer14; 21.2% vs 0%, p-value = 1.694e-11 for p.Trp304Ter) (Table 3). All identified variants except for p.Arg594Gln were absent from the Egypt Control cohort, where p.Arg594Gln was present in only 0.25% (1/400) of the cohort in the heterozygous form.

**Table 3:**
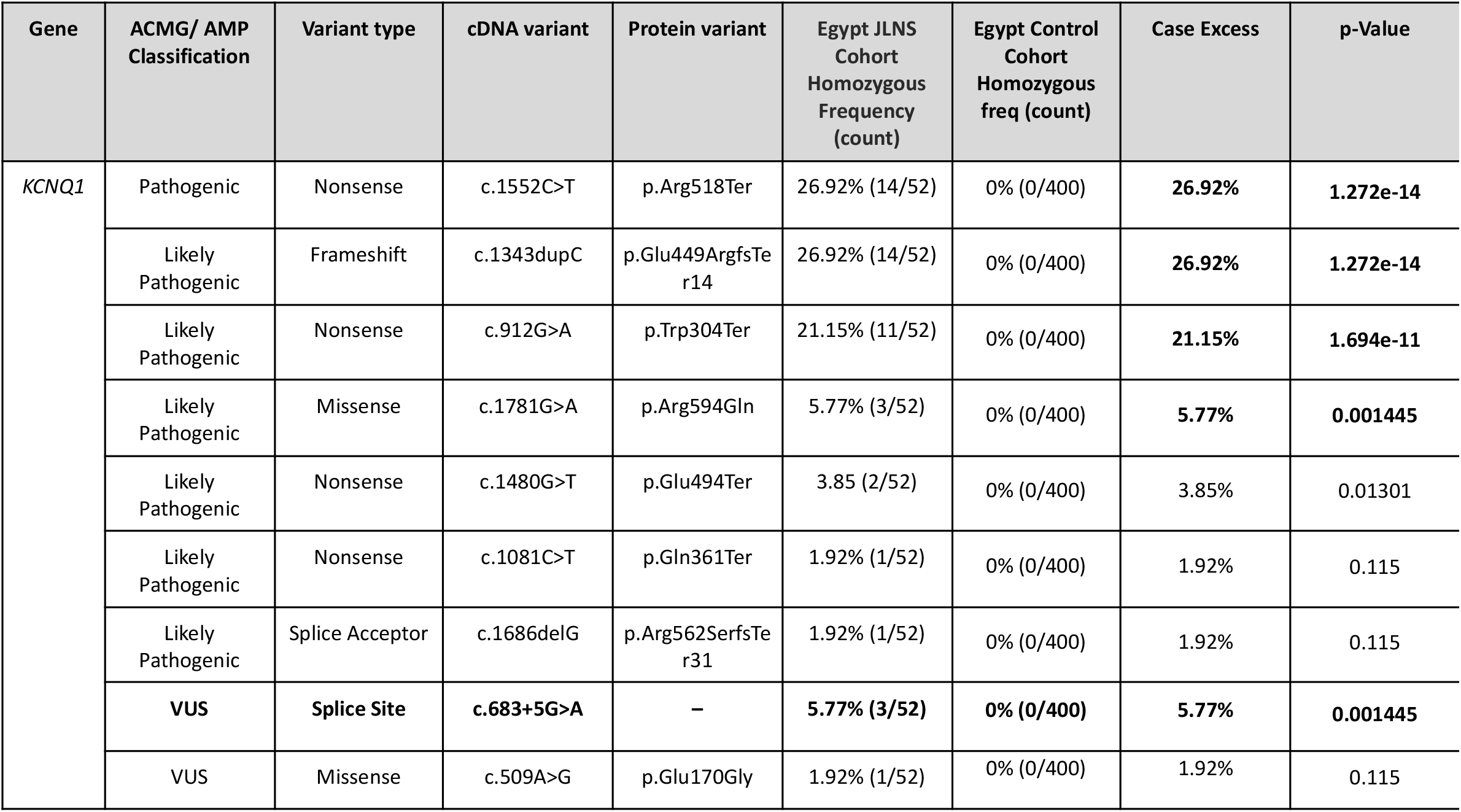
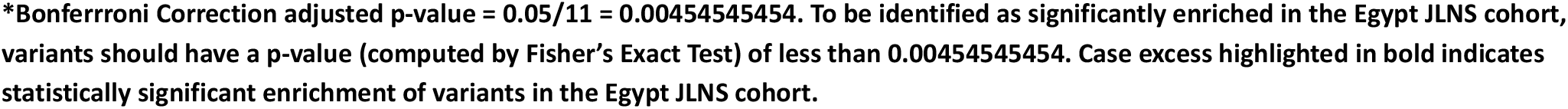
Summary of rare homozygous *KCNQ1* and *KCNE1* genetic variants identified in the Egypt JLNS Cohort.

All identified homozygous variants were absent from gnomAD (v2.1.1), underscoring their potential pathogenicity. Of these homozygous variants, only two (*KCNQ1*: p.Arg518Ter and p.Arg594Gln) were found in gnomAD (0.0001156 and 0.00001123, respectively in European (Non-Finnish) populations) in heterozygosity, suggesting that a double-hit is required to cause disease.

Additionally, three variants (c.912G>A (p.Trp304Ter), c.1686delG, and p.Glu170Gly) are likely novel variants due to their absence from gnomAD, the Greater Middle East Variome database, and Clinvar. Moreover, the novel *KCNQ1* c.912G>A variant was reported in 11 patients from 10 unique families, suggesting that it could be a North African-specific founder.

### Distribution of rare variants across the KCNQ1 protein and their cardiac phenotype

Since the majority of sequenced patients (51/52 patients) harbored homozygous variants in *KCNQ1*, the distribution of rare variants across the *KCNQ1* protein was investigated. The majority of variants (35/51) were distributed across the cytosolic carboxyl terminus (C-terminal) of the KCNQ1 protein (Figure 3).

**Figure 3:**
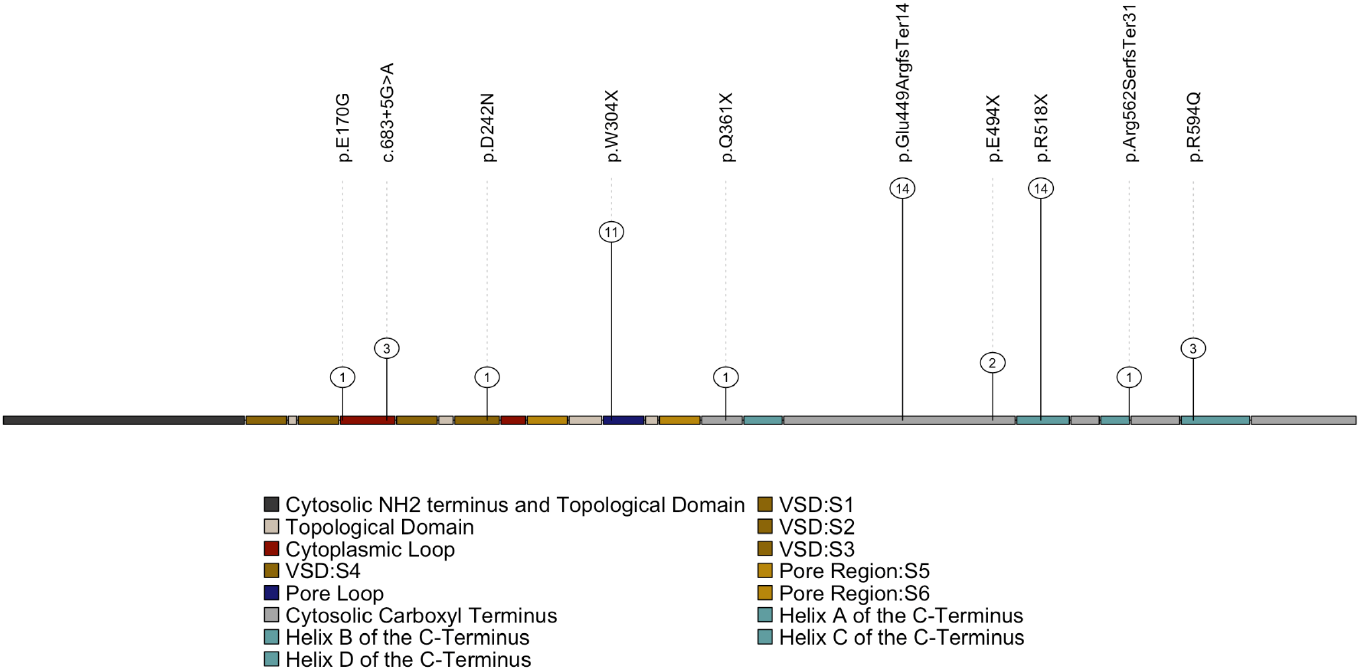
Distribution of the identified variants ccross the KCNQ1 Protein. The numbers in the nodes represent the proportion of individuals with the identified homozygous variants.

Subsequently, we compared the cardiac phenotype of carriers of variants located in the c-terminal of the KCNQ1 protein versus that of carriers of variants located across domains of the KCNQ1 protein within the Egypt JLNS cases (Table 4). None of the cardiac parameters were significantly different between the carriers of variants located in the c-terminal of the KCNQ1 protein and carriers of variants located across domains of the KCNQ1 protein (Table 4).

**Table 4:**
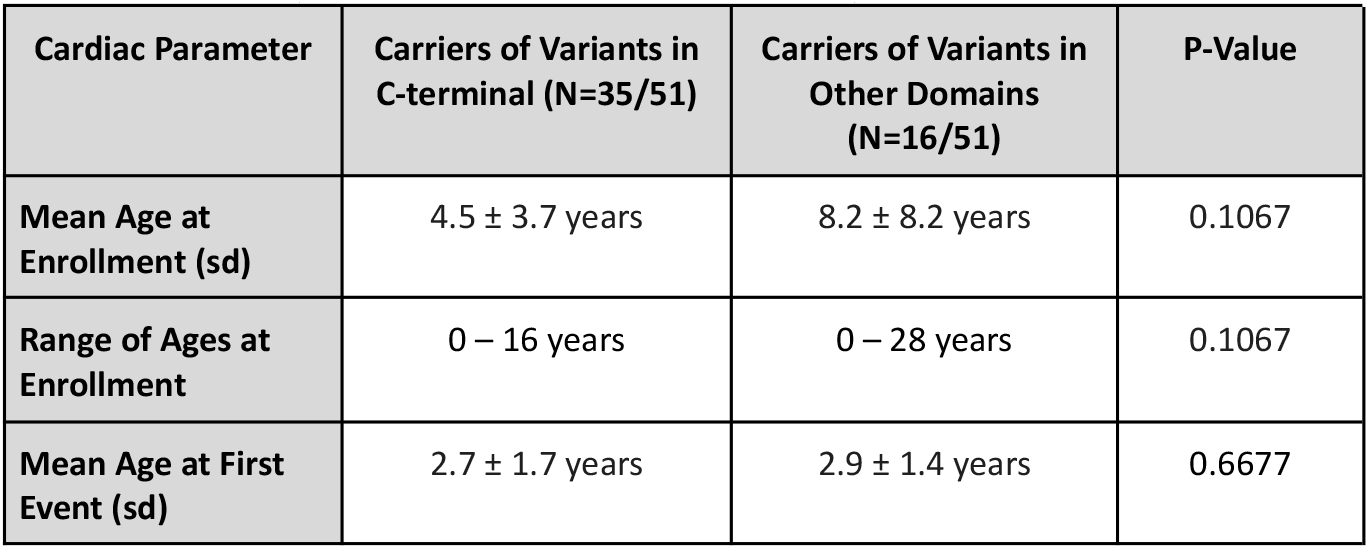
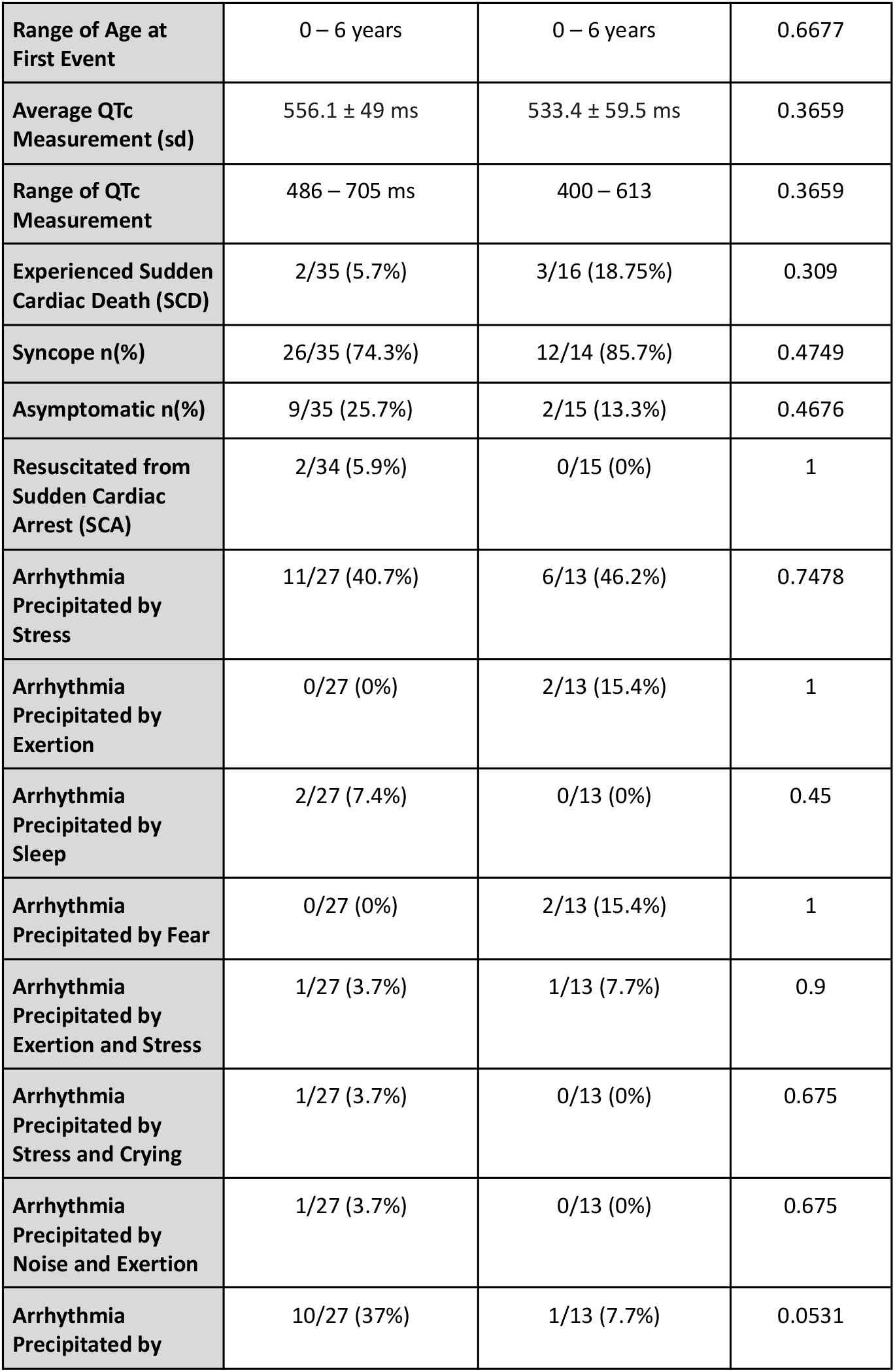
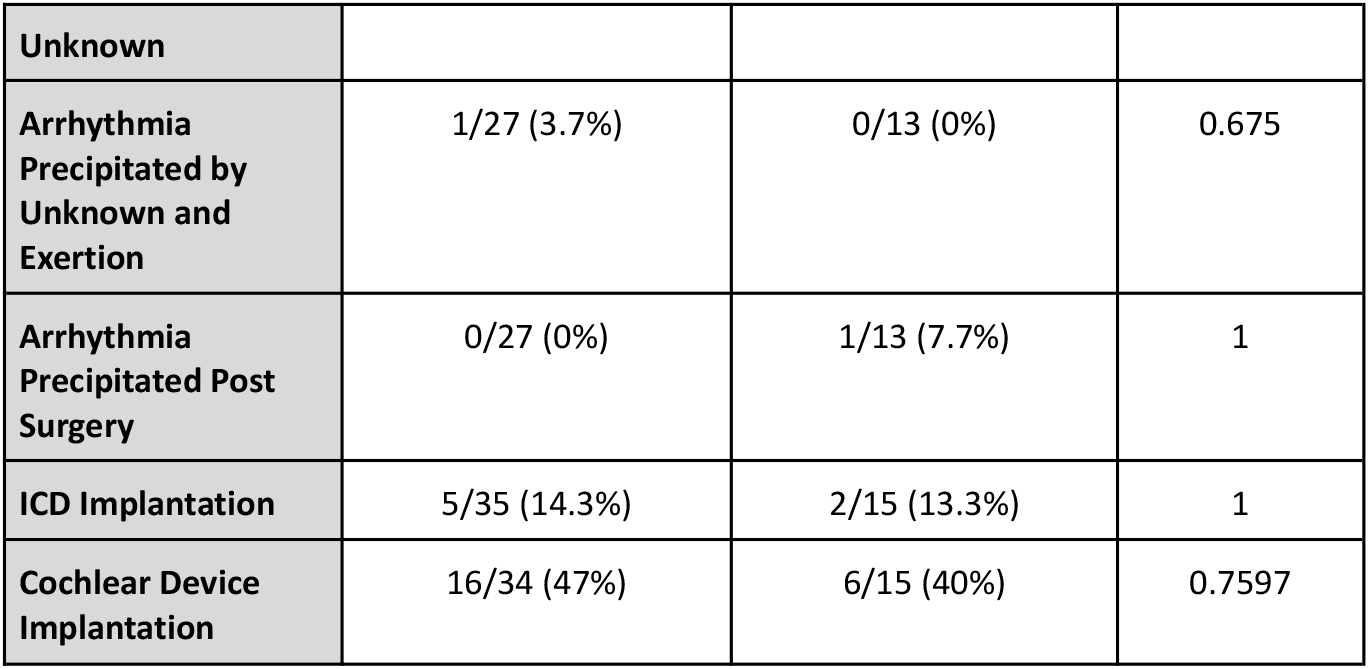
Comparison of Cardiac Phenotype between Carriers of Variants Located in the C-Terminal of KCNQ1 versus Other Domains of the KCNQ1 Protein.

### Clinical Characteristics of Carriers of Truncating vs Nontruncating Variants

The majority (90.4%) of identified variants in *KCNQ1* or *KCNE1* were truncating (Table 3). In fact, only 5 patients (9.6%) carried a non-truncating (i.e. missense) variant versus 47 patients (90.4%) who carried a truncating (i.e. frameshift, nonsense, splice acceptor, or splice site) variant (Table 3). We compared the clinical characteristics between the carriers of the two types of variants and found no significant differences in any cardiac parameter (Supplementary Table 3).

### The Interplay of Homozygosity and Consanguinity

Consanguinity was self-reported in 77.5% (31/40) of families for which complete family history data were available, indicating a substantial prevalence of familial relatedness within the studied cohort. This high rate of consanguinity likely contributes to the observed enrichment of homozygous variants in the cohort, where all 44 unrelated patients from unique families harbored homozygous variants in either *KCNQ1* or *KCNE1*. Specifically, all 44 unrelated patients from unrelated families were found to carry homozygous pathogenic variants in either *KCNQ1* or *KCNE1;* whereas, all parents harbored heterozygous variants and were confirmed to be healthy through electrophysiological evaluation.

## Discussion

This study represents the largest case-control analysis of JLNS in the MENA region to date. We also report the highest proportion of consanguineous JLNS families, accounting for 76.2% of families. In comparison, the largest JLNS cohort published to date, reported by Schwartz et al (2006), found consanguinity in only 35% of JLNS families and 41% of sequenced families^9^. The high rate of consanguinity is a unique aspect of our study, whereas the two other large JLNS studies reported compound heterozygous variants in 33% and almost 7% of the cohort, respectively^9,38^. This reinforces the strong interplay between consanguinity and homozygosity.

In regards to the characteristics of the cohort, there was a higher predominance of females compared to males in our cohort. This is consistent with the proportion of females to males described in the recently published study by Charles Morgat et al (2024)^38^. We compared the electrocardiographic phenotype across genders and disease groups among JLNS patients and at least one of their parents Similar to previous studies, no differences were observed in the QTc measurements between the sexes of patients^9,13^. Similarly, no differences were observed in the QTc measurements between sex groups of parents of JLNS patients. However, significant differences were observed between JLNS patients and their parents, regardless of the sex group. This data further proves that gender groups do not impact the QTc measurements across JLNS patients and parents of JLNS patients. Additionally, when comparing symptomatic and asymptomatic JLNS patients, no significant differences in QTc measurements were found, which is consistent with the literature^9^.

In addition to these gender-specific phenotypic observations, we examined the diagnostic yield in our cohort. The analysis showed that the yield was 100% among all Egypt JLNS patients which is in congruence with previous studies^9,38^. Additionally, the proportion of patients in the Egypt JLNS cohort, who harbored variants in *KCNQ1* (98%) or *KCNE1* (2%), was similar to those of the JLNS patients in the Schwartz et al (2006) study (*KCNQ1* variants (95%) and *KCNE1* variants (5%))^9^. To date, no JLNS cohort study has publicly shared the distribution of the JLNS patients carrying genetic variants in *KCNQ1* or *KCNE1*, according to the ACMG/AMP variant classification guidelines. Likewise, to date, no JLNS cohort study has investigated the enrichment of specific variants in the cohort against ancestry matched controls. Of the three significantly enriched variants in our Egypt JLNS cohort (p.Arg518Ter, p.Glu449ArgfsTer14, and p.Trp304Ter), p.Arg518Ter was also found to be enriched in the Swedish JLNS cases^13,39^. However, genealogical and haplotype analyses of an aggregate of both cohorts are needed to confirm that the p.Arg518Ter is indeed a Swedish common founder variant rather than an Egyptian one, given its enrichment in the Egypt JLNS cases. Moreover, it is worth noting that the *KCNQ1* variant p.Trp304Ter identified in our cohort has a different variant (c.912G>A) than the one reported in the ClinVar database (c.911G>A). Hence, it is highly probable that the variant identified in our cohort is most likely an Egyptian founder mutation.

Furthermore, upon the investigation of the localization of the variants on the KCNQ1 protein, we found that the majority of variants are localized in the cytosolic carboxylic terminal (c-terminal) of the protein. In their recent study, Morgat and Colleagues (2024) have mentioned that variants localized in the c-terminal region of the KCNQ1 protein are reportedly associated with a lower risk of arrhythmic events^38^. Therefore, we compared the cardiac parameters across carriers of variants located in the c-terminal region and carriers of variants located in other domains of the KCNQ1 protein. Our results showed no significant differences between the carriers of both variant locations.

It is worth noting that in many inherited cardiac diseases, truncating variants are typically associated with more severe clinical manifestations. However, this pattern does not appear to apply to JLNS. Both our study and the work of Schwartz and colleagues (2006) have demonstrated that there are no significant differences in electrocardiographic phenotypes or other arrhythmic manifestations between carriers of truncating versus non-truncating variants^9^. However, a larger cohort size is needed to further validate these findings.

Although this study represents one of the largest JLNS cohort studies, the relatively modest sample size (n=57) limits statistical power to detect subtle genotype-phenotype associations and to perform detailed subgroup analyses. Additionally, data on consanguinity was self-reported, which may have introduced reporting bias. Furthermore, the genetic analyses of this study focused solely on *KCNQ1* and *KCNE1* genes, which might have potentially overlooked rare variants in other genes or non-coding regions that might contribute to JLNS.

In conclusion, variants discovered in this underrepresented and highly consanguineous population contribute to the expanding mutational spectrum of JLNS and highlight ancestry-specific genetic determinants of disease. Further expansion of this cohort, coupled with in-depth functional and molecular investigations, will be pivotal in uncovering novel genetic and mechanistic insights, ultimately advancing the global understanding, diagnosis, and management of inherited arrhythmia syndromes.

## Data Availability

All data produced in the present work are contained in the manuscript and available upon reasonable request to the authors.

