## Supplementary Material for "The mutational spectrum of Jervell and Lange-Nielsen syndrome: insights from highly consanguineous families"

**Supplementary Table 1: Results of the Tukey HSD Test across the different comparison groups.**

| | Difference in Means | Lower Bound | Upper Bound | Adjusted P-Value | Significance (Adjusted P-Value < 0.05) |
|---|---|---|---|---|---|
| JLNS Parent_Male-JLNS Parent_Female | -23.953333 | -55.834153 | 7.92748663 | 0.20945199 | Not Significant |
| JLNS Patient_Female-JLNS Parent_Female | 106.472917 | 76.554473 | 136.39136 | 6.15063555642337e-14 | *** |
| JLNS Patient_Male-JLNS Parent_Female | 109.366667 | 77.4858467 | 141.247487 | 1.17239551400417e-13 | *** |
| JLNS Patient_Female-JLNS Parent_Male | 130.42625 | 99.0015454 | 161.850955 | 3.70814490224802e-14 | *** |
| JLNS Patient_Male-JLNS Parent_Male | 133.32 | 100.02157 | 166.61843 | 4.39648317751562e-14 | *** |
| JLNS Patient_Male-JLNS Patient_Female | 2.89375 | -28.530955 | 34.3184546 | 0.99509209 | Not Significant |

*** represent p-values < 0.001.

**Supplementary Table 2: Details of the electrocardiographic phenotype of JLNS patients versus their parents, categorized by gender.**

| | Female JLNS Patients | Male JLNS Patients | Female Parents of JLNS patients | Male Parents of JLNS patients |
|---|---|---|---|---|
| **Average QTc Measurement (sd)** | 541.9 ± 58.9 ms | 544.8 ± 52.9 ms | 435.4 ± 31.1 ms | 411.5 ± 26.7 ms |
| **Range of QTc Measurement** | 400 – 705 ms | 449 – 656 ms | 348 – 480 ms | 361 – 465 ms |
| **Median QTc Measurement** | 543 ms | 540 ms | 441 ms | 408 ms |

**Supplementary Table 3: Comparison of Clinical Parameters between Truncating and Non-Truncating Variant Carriers**

| Clinical Parameter | Carriers of Truncating Variants | Carriers of Nontruncating Variants | P-Value |
|---|---|---|---|
| **Mean Age at Enrollment (sd)** | 6 ± 5.9 years | 2.6 ± 1.9 years | 0.09491 |
| **Range of Ages at Enrollment** | 0 – 28 years | 0 – 5 years | 0.09491 |
| **Mean Age at First Event (sd)** | 2.8 ± 1.6 years | 2 ± 1 years | 0.2905 |
| **Range of Age at First Event** | 0 – 6 years | 1 – 3 years | 0.2905 |
| **Average QTc Measurement** | 550.1 ± 52.7 ms | 529.6 ± 56.7 ms | 0.4104 |
| **Range of QTc Measurement** | 400 – 705 ms | 462 – 613 ms | 0.4104 |
| **Experienced Sudden Cardiac Death (SCD)** | 4/47 (8.5%) | 1/5 (20%) | 0.4098 |
| **Syncope n(%)** | 35/45 (77.8%) | 3/5 (60%) | 0.5819 |
| **Asymptomatic n(%)** | 10/46 (21.7%) | 2/5 | 0.5798 |
| **Resuscitated from Sudden Cardiac Arrest (SCA)** | 2/45 (4.4%) | 0/5 | 1 |
| **Arrhythmia Precipitated by Stress** | 14/37 (37.8%) | 3/3 (100%) | 0.06882591 |
| **Arrhythmia Precipitated by Exertion** | 2/37 (5.4%) | 0/3 (0%) | 1 |
| **Arrhythmia Precipitated by Sleep** | 2/37 (5.4%) | 0/3 (0%) | 1 |

| | | | |
|---|---|---|---|
| **Arrhythmia Precipitated by Fear** | 2/37 (5.4%) | 0/3 (0%) | 1 |
| **Arrhythmia Precipitated by Exertion and Stress** | 2/37 (5.4%) | 0/3 (0%) | 1 |
| **Arrhythmia Precipitated by Stress and Crying** | 1/37 (2.7%) | 0/3 (0%) | 1 |
| **Arrhythmia Precipitated by Noise and Exertion** | 1/37 (2.7%) | 0/3 (0%) | 1 |
| **Arrhythmia Precipitated by Unknown** | 11/37 (29.7%) | 0/3 (0%) | 1 |
| **Arrhythmia Precipitated Post Surgery** | 1/37 (2.7%) | 0/3 (0%) | 1 |
| **ICD Implantation** | 6/46 (13%) | 1/5 (20%) | 0.5377 |
| **Cochlear Device Implantation** | 22/45 (48.9%) | 1/5 (20%) | 0.3573 |
